# Alzheimer’s disease as a form of in situ diabetes: A systematic review and meta-analysis of therapeutic approaches using intranasal insulin treatment

**DOI:** 10.1101/2024.06.21.24309323

**Authors:** Luís Jesuíno de Oliveira Andrade, Gabriela Correia Matos de Oliveira, Luís Matos de Oliveira

**Affiliations:** Departamento de Saúde da Universidade Estadual de Santa Cruz - Ilhéus - Bahia - Brasil; Médica pela Faculdade de Medicina UniFTC - Salvador - Bahia - Brasil; Escola Bahiana de Medicina e Saúde Pública - Salvador - Bahia - Brasil

**Keywords:** Alzheimer’s disease, Intranasal insulin, Systematic review, Meta-analysis

## Abstract

**Introduction:** Alzheimer’s disease (AD) is a progressive neurodegenerative disorder that is characterized by the loss of memory, language, and other cognitive functions. Increasing evidence suggests that AD shares pathophysiological similarities with type 2 diabetes, leading to the concept of AD as “diabetes in situ” within the brain. Intranasal insulin (INI) for AD has emerged as a promising therapeutic approach due to its ability to directly target the brain and modulate insulin signaling pathways.

**Objective:** To evaluate the efficacy and safety of INI therapy for AD through a systematic review and meta-analysis of randomized clinical trials.

**Method:** A comprehensive search across electronic databases, including PubMed, Web of Science, Scopus, and Embase, was conducted to identify relevant studies published up to June 2024. Studies were included if they met the following criteria: original research articles published in peer-reviewed journals; focused on humans; investigated the therapeutic effects of INI administration on cognitive impairment associated with AD or diabetes; reported quantitative data on cognitive outcomes, biomarkers, or pathological markers relevant to AD or diabetes. For studies with available data, a meta-analysis was conducted to quantitatively synthesize the effects of INI on cognitive outcomes. METAANALYSISONLINE (https://metaanalysisonline.com/), an online statistical tool, was employed to conduct the meta-analysis and generate forest plots and funnel plots.

**Results:** A total of 647 articles were identified through electronic database searches using predefined search terms, and eight studies met the inclusion criteria and were selected for data extraction and analysis. Based on the analysis performed using random effects model with Mantel-Haenszel method to compare the odds ratio, the overall odds ratio was 3.75 with a 95% confidence interval of 1.49 - 9.4. The test for overall effect shows a significance at p<0.05. The The I^2^ value indicates that 85.5% of the variability among studies arises from heterogeneity rather than random chance.

**Conclusion:** While the data is not yet definitive enough to establish INI as a definitive treatment for AD, the accumulating evidence supporting its safety, efficacy, and reduced systemic side effects strongly suggests that INI is associated with an overall enhancement of global cognition.

## INTRODUCTION

Alzheimer’s disease (AD) is a progressive neurodegenerative disorder that is characterized by the loss of memory, language, and other cognitive functions. It is the most common cause of dementia, accounting for 60-80% of cases.^1^ The pathophysiology of AD is complex and not fully understood. However, two key neuropathological hallmarks are consistently observed in AD brains: amyloid plaques and neurofibrillary tangles. Amyloid plaques are composed of a protein called amyloid-beta (Aβ), while neurofibrillary tangles are composed of a protein called tau. These abnormal protein aggregates are thought to disrupt normal brain function and lead to neuronal death.^2^ There is no cure for AD, and current treatments are only able to temporarily manage the symptoms. However, there is a great deal of research underway to develop new treatments and therapies for AD. Some promising approaches include targeting Aβ and tau aggregation, modulating neuroinflammation, and enhancing neurogenesis.^3^ Despite the challenges, there is hope that AD will one day be a preventable and treatable disease.

Brain insulin resistance (BIR) is a condition characterized by the impaired ability of brain cells to respond to insulin. While insulin is primarily known for its role in peripheral glucose homeostasis, it also plays a crucial role in brain function, including memory, cognition, and learning.^4^ BIR has been implicated in the pathogenesis of various neurodegenerative diseases, including AD.^5^ In AD, BIR is associated with increased amyloid plaque deposition and tau neurofibrillary tangles, two hallmark neuropathological features of the disease.^6^ The mechanisms underlying BIR in the brain are complex and not fully understood. However, several factors are thought to contribute, including chronic hyperglycemia, obesity, and inflammation.^7^ Current therapeutic approaches for BIR in the brain are limited.

Intranasal insulin (INI) for AD has emerged as a promising therapeutic approach due to its ability to directly target the brain and modulate insulin signaling pathways. INI offers several advantages over systemic insulin administration. Bypassing the blood-brain barrier allows INI to deliver insulin directly to brain cells, maximizing its therapeutic effect while minimizing systemic side effects.^8^ The mechanisms underlying INI’s beneficial effects in AD are multifaceted. The INI promotes glucose uptake and energy metabolism in neurons, modulates neuroinflammation, and stimulates neurogenesis, which may help counteract neuronal loss in AD.^9^

Increasing evidence suggests that AD shares pathophysiological similarities with type 2 diabetes, leading to the concept of AD as “diabetes in situ” within the brain. In this way, the administration of INI, by crossing the blood-brain barrier, allows for the delivery of insulin directly to brain cells, maximizing its effect on the brain and with rare systemic side effects. The aim of this study is to evaluate the efficacy of INI therapy for AD through a systematic review and meta-analysis of randomized clinical trials (RCTs).

## METHODOLOGY

### Study Design, Protocol and registration

To ensure transparency and methodological rigor, our systematic review protocol was prospectively registered on the International Prospective Register of Systematic Reviews (PROSPERO): CRD42024 560578. This public record, accessible at (https://www.crd.york.ac.uk/prospero/%23), outlines the planned search strategy, inclusion/exclusion criteria, and data extraction methods employed in the review. Furthermore, we adhered to the established Preferred Reporting Items for Systematic Reviews and Meta-Analyses (PRISMA) guidelines throughout the review process,^10^ and included RCTs that comprehensively investigated the literature on the potential efficacy of INI for AD.

### Literature Search Strategy

A comprehensive search across electronic databases, including PubMed, Web of Science, Scopus, and Embase, was conducted to identify relevant studies published up to June 2024. The search strategy employed a combination of controlled vocabulary and free-text terms related to AD and INI therapy. Boolean operators (AND, OR, NOT) were utilized to refine the search strategy and ensure the retrieval of relevant studies. Specific search terms and variations included: (“alzheimer disease”[MeSH Terms] OR (“alzheimer”[All Fields] AND “disease”[All Fields]) OR “alzheimer disease”[All Fields]) AND (intranasal[All Fields] AND (“insulin”[MeSH Terms] OR “insulin”[All Fields])). To ensure a comprehensive search, references of included studies and relevant review articles were also examined.

### Inclusion and Exclusion Criteria

Studies were included if they met the following criteria: Original research articles published in peer-reviewed journals; Focused on humans ; Investigated the therapeutic effects of intranasal insulin administration on cognitive impairment associated with AD or diabetes; Reported quantitative data on cognitive outcomes, biomarkers, or pathological markers relevant to AD or diabetes.

Studies were excluded if they were review articles, editorials, commentaries, case reports, or abstracts without sufficient data. Studies that examined only peripheral metabolic effects without evaluating cognitive or neurological outcomes were also excluded.

### Data Extraction and Synthesis - Study quality assessment

To ensure methodological rigor and transparency, we employed the Cochrane risk-of-bias tool version 2 (ROB2) specifically designed for evaluating randomized controlled trials.^11^ This standardized tool facilitates a systematic assessment of potential bias across critical domains that can influence study outcomes. These domains encompass: the generation of random sequences to prevent allocation predictability; the implementation of allocation concealment to mask treatment assignment until intervention initiation; the blinding of participants, personnel administering interventions, and outcome assessors to minimize performance and detection bias; the extent of missing outcome data to evaluate the potential for attrition bias; the selective reporting of pre-specified outcomes to safeguard against reporting bias; and the exploration of any other potential sources of bias not captured within the aforementioned domains. Through this comprehensive evaluation using ROB2, the risk of bias impacting the authors’ conclusions was categorized as either “low risk,” “some concerns,” or “high risk”.

Relevant data from included studies were extracted using a standardized data extraction form, including study characteristics (authors, publication year, study design, sample size), participant demographics, details of intranasal insulin administration (dose, frequency, duration), cognitive assessment tools, and main outcomes.

For studies with available data, a meta-analysis was conducted to quantitatively synthesize the effects of INI on cognitive outcomes. Standardized mean differences and 95% confidence intervals (CIs) were calculated for continuous outcomes, while odds ratios (ORs) and 95% CIs were calculated for dichotomous outcomes. Random-effects models were employed to account for potential heterogeneity between studies. Subgroup analyses were conducted based on the type of population (AD or diabetes) and the specific cognitive domain assessed.

### Statistical Analysis

METAANALYSISONLINE (https://metaanalysisonline.com/),^12^ an online statistical tool, was employed to conduct the meta-analysis and generate forest plots and funnel plots. Heterogeneity among studies was assessed using the Q statistic and the I^2^ index, with I^2^ values of 25%, 50%, and 75% representing low, moderate, and high heterogeneity, respectively. Publication bias was evaluated through visual inspection of funnel plots and Egger’s regression test. Sensitivity analyses were conducted to assess the robustness of the results, excluding studies with high risk of bias or outliers.

### Ethical Considerations

This systematic review and meta-analysis obeys the ethical principles of research integrity and transparency. Data was extracted and analyzed objectively without any conflicts of interest. The findings are presented in a clear and concise manner, adhering to scientific standards.

## RESULTS

A comprehensive overview of the search strategy is presented in the PRISMA flowchart (Figure 1), highlighting the stepwise process of study retrieval and inclusion for this systematic review. A total of 647 articles were identified through electronic database searches using predefined search terms. Upon removal of duplicates, review articles, editorials, commentaries, case reports, or abstracts without sufficient data, 193 articles underwent abstract screening and full-text review, with selection based on the investigation of the association between BIR and AD. Ultimately, eight studies met the inclusion criteria and were selected for data extraction and analysis.

**Figure 1.**
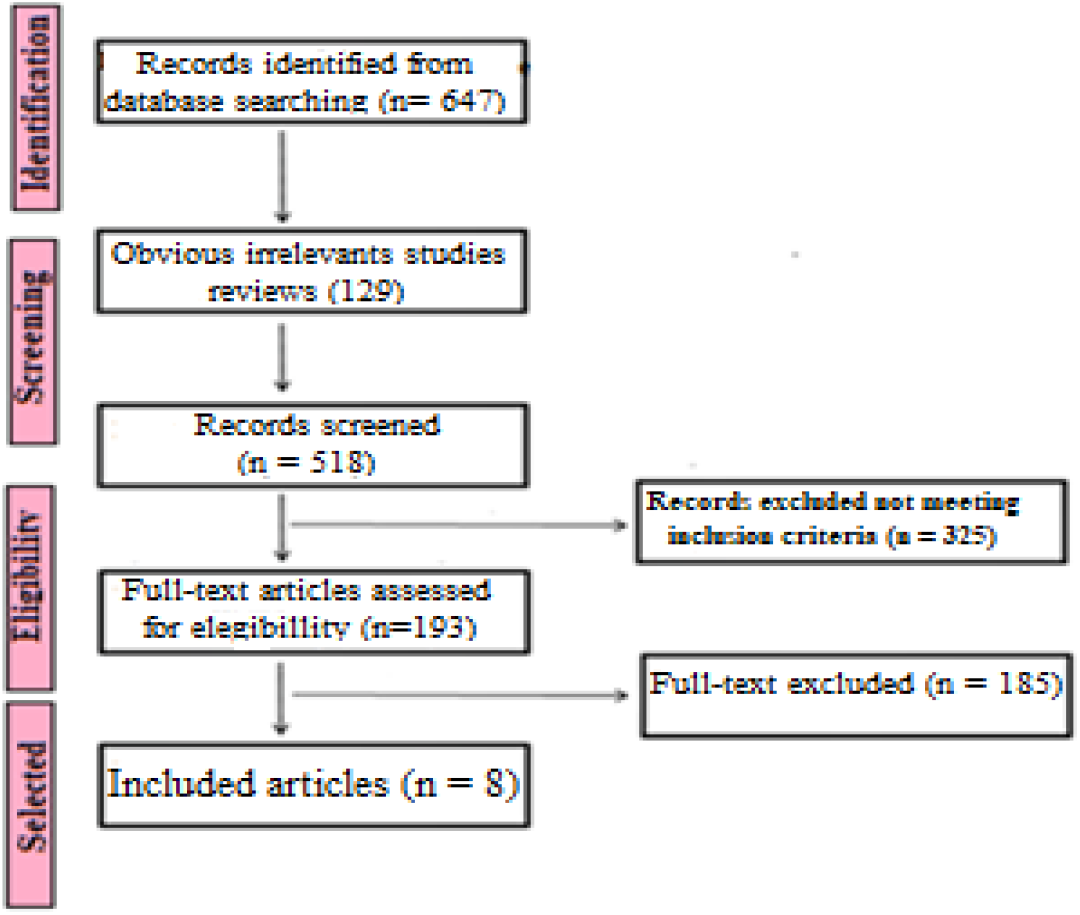
Flowchart of the selection process for the 8 studies included.

All selected studies explored the link between INI and AD. Of the eight studies that met the inclusion criteria, all were case-control studies. The studies were conducted across different countries, with sample sizes ranging from 35 to 529 participants.

### Relevant Characteristics of the Included Studies

- ***Reference***: Craft S, et al.^13^; Type of study: Case-control study; Objective: “To examine the feasibility, safety, and efficacy of intranasal insulin for the treatment of persons with mild cognitive impairment (MCI) and AD dementia in a phase 2/3 multisite clinical trial”; Participants: A total of 289 cases and 240 controls; Conclusion: “In this study, no cognitive or functional benefits were observed with intranasal insulin treatment over a 12-month period among the primary intention-to-treat cohort”.
- ***Reference***: Kellar D, et al.^14^; Type of study: Case-control study; Objective: “To assess the effects of INI on cerebral spinal fluid markers of inflammation, immune function, and vascular function and their associations with clinical markers of AD progression”; Participants: A total of 24 cases and 25 controls; Conclusion: “INI treatment altered the typical progression of markers of inflammation and immune function seen in AD, suggesting that INI may promote a compensatory immune response associated with therapeutic benefit”.
- ***Reference***: Rosenbloom M, et al.^15^; Type of study: Case-control study; Objective: “To evaluate the safety and efficacy of rapid-acting intranasal glulisine in subjects with amnestic MCI or mild probable AD”; Participants: A total of 19 cases and 16 controls; Conclusion: “There were no enhancing effects of intranasal glulisine on cognition, function, or mood, but the ability to detect significance was limited by the number of subjects successfully enrolled and the study duration”.
- ***Reference***: Kellar D, et al.^16^; Type of study: Case-control study; Objective: “ To assess the effects of intranasally administered insulin on white matter health and its association with cognition and cerebral spinal fluid biomarker profiles in adults with MCI or AD in secondary analyses from a prior phase 2 clinical trial (NCT01767909); Participants: A total of 19 cases and 16 controls; Conclusion: “Intranasal insulin treatment for 12 months reduced white matter hyperintensity volume progression and supports insulin’s potential as a therapeutic option for AD”.
- ***Reference***: Craft S, et al.^17^; Type of study: Case-control study; Objective: “To examine the effects of intranasal insulin administration on cognition, function, cerebral glucose metabolism, and cerebrospinal fluid biomarkers in adults with amnestic MCI or AD”; Participants: A total of 104 cases and 104 controls; Conclusion: “The results support longer trials of intranasal insulin therapy for patients with amnestic MCI and patients with AD”.
- ***Reference***: Craft S, et al.^18^; Type of study: Case-control study; Objective: “To determine whether four months of treatment with intranasal insulin detemir or regular insulin improves cognition, daily functioning, and AD biomarkers for adults with MCI or AD”; Participants: A total of 24 cases and 13 controls; Conclusion: “Future research is warranted to examine the mechanistic basis of treatment differences, and to further assess the efficacy and safety of intranasal insulin”.
- ***Reference***: Claxton A, et al.^19^; Type of study: Case-control study; Objective: “To examined the safety profile and efficacy of two doses of insulin detemir for treatment of adults diagnosed with AD or amnestic MCI compared with placebo”; 9Participants: A total of 60 cases and 20 controls; Conclusion: “Daily treatment with 40 IU insulin detemir modulated cognition for adults with AD or amnestic mild cognitive, with APOE-related differences in treatment response for the primary memory composite”.
- ***Reference***: Claxton A, et al.^20^; Type of study: Case-control study; Objective: “To evaluate sex and ApoE genotype differences in treatment response to two doses of intranasal insulin in adults with MCI or AD”; Participants: A total of 74 cases and 30 controls; Conclusion: “There has been evidence that intranasal insulin is a safe and effective treatment for the memory loss associated with MCI and AD. The current paper suggests that treatment response may vary by sex and ApoE ε4 carriage”.

Table 1 provides an overview of the included studies.

**Tabela 1.** General Characteristics of the Studies Included.

| Author | Year | Study type | Total Case / positive outcomes | Total control / positive outcomes |
| --- | --- | --- | --- | --- |
| Craft S, et al.<sup>13</sup> | 2021 | case-control | 289/240 | 240/49 |
| Kellar D, et al.<sup>14</sup> | 2022 | case-control | 24/18 | 25/4 |
| Rosenbloom M, et al.<sup>15</sup> | 2021 | case-control | 19/2 | 16/2 |
| Kellar D, et al.<sup>16</sup> | 2021 | case-control | 19/15 | 16/12 |
| Craft S, et al.<sup>17</sup> | 2012 | case-control | 104/66 | 104/42 |
| Craft S, et al.<sup>18</sup> | 2017 | case-control | 24/19 | 13/8 |
| Claxton A, et al.<sup>19</sup> | 2015 | case-control | 60/24 | 20/4 |
| Claxton A, et al.<sup>20</sup> | 2013 | case-control | 74/25 | 30/4 |

### Quality assessment

Systematic bias or error can be defined as any tendency in data collection, analysis, interpretation, publication, or review that leads to conclusions that systematically deviate from the truth. Despite the gold standard nature of RCTs in human research, this type of study is highly susceptible to bias, whether due to investigator arbitrariness in sample selection and measurement of analyzed variables, or due to the difficulty in controlling other factors that may influence clinical outcomes.

The studies were subjected to a bias risk assessment using the Cochrane Collaboration Network’s RoB (Risk of Bias) tool. Following the assessment, it was determined that all studies presented low risk. However, the risk associated with “random sequence generation” and overall RoB remained unclear in all studies.

Additionally, the RoBs of two studies were considered unclear due to uncertainties related to “participant blinding” and/or “allocation concealment.” The detailed RoB assessments are presented in Figures 2 and 3. The crossover study by Rosenbloom et al.^15^ was identified as low risk based on its study design, as randomization of the treatment sequence did not produce subsequent treatment effects.

**Figure 2.**
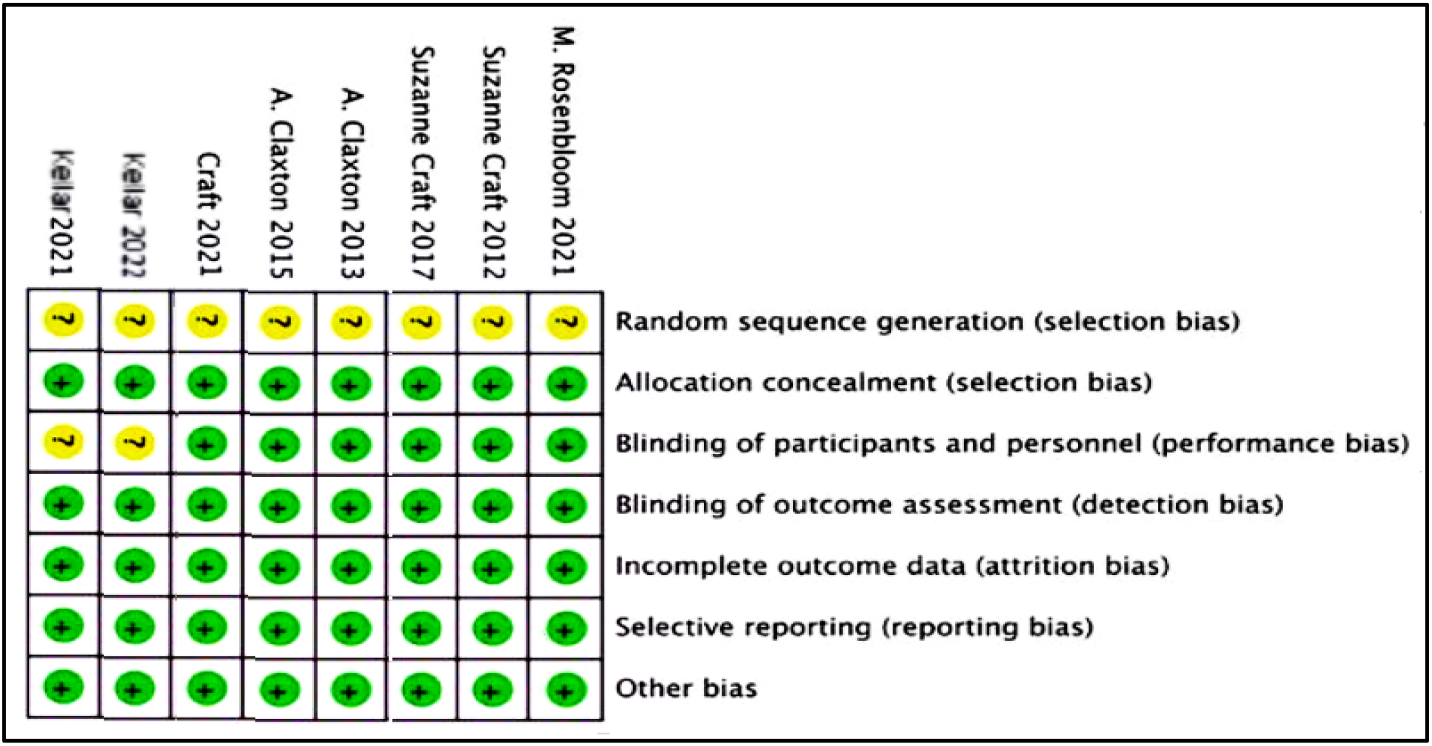
Risk of bias summary

**Figure 3.**
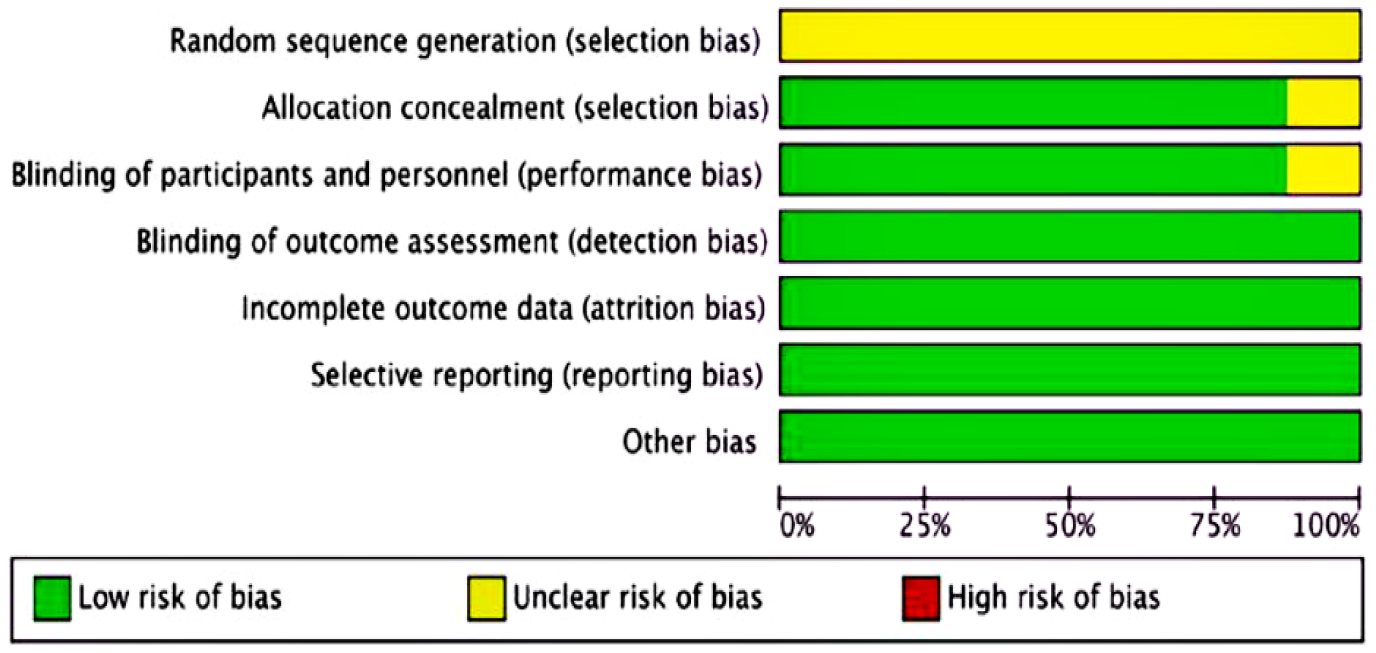
Risk of bias graph presented as percentage of the included studies

### Meta-Analysis of Intranasal Insulin for Alzheimer’s disease

This investigation employed a rigorous meta-analysis approach to assess the potential benefits of INS for AD. A comprehensive search strategy was implemented, encompassing all pertinent association studies published up to June 2024. To ensure inclusivity, we conducted a meticulous search across major electronic databases, including PubMed, Web of Science, Scopus, and Embase.

Our meta-analysis yielded a total of eight original case-control studies examining the therapeutic response to intranasal insulin in AD. These studies collectively encompassed a substantial sample size of 613 individuals.

A total of eight studies were analyzed, including 613 participants in the experimental cohort and 464 participants in the control cohort. Based on the analysis performed using random effects model with Mantel-Haenszel method to compare the odds ratio, there is a statistical difference between the two cohorts, the overall odds ratio is 3.75 with a 95% confidence interval of 1.49 - 9.4. The test for overall effect shows a significance at p<0.05. A significant heterogeneity was detected (0), suggesting inconsistent effects in magnitude and/or direction. The The I^2^ value indicates that 85.5% of the variability among studies arises from heterogeneity rather than random chance (Figure 4).

**Figure 4.**
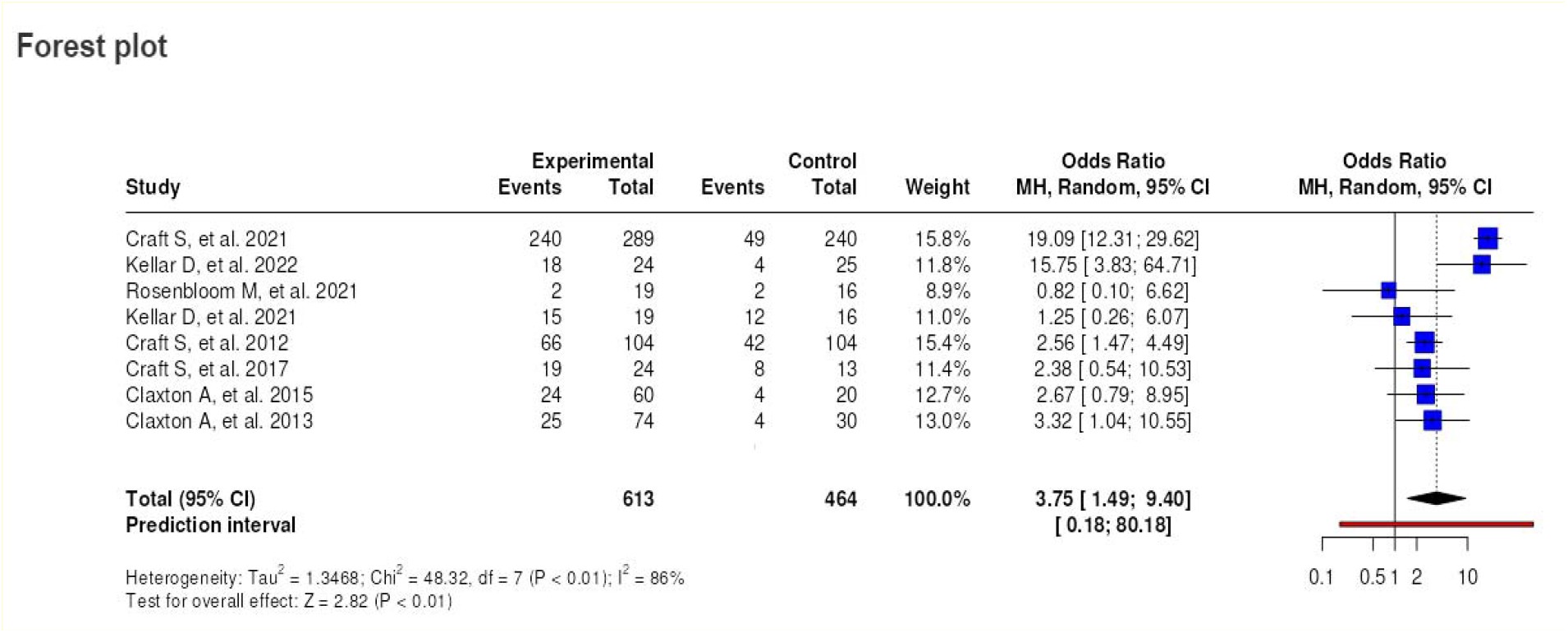
Comparative assessment of intranasal insulin (INI) versus placebo on global cognitive function in the overall study population

The funnel plot does not indicate a potential publication bias. The Eggers’ test does not support the presence of funnel plot asymmetry (intercept: -2.77, 95% CI:-5.98 - 0.44, t: -1.689, p-value: 0.142) (Figure 5).

**Figure 5.**
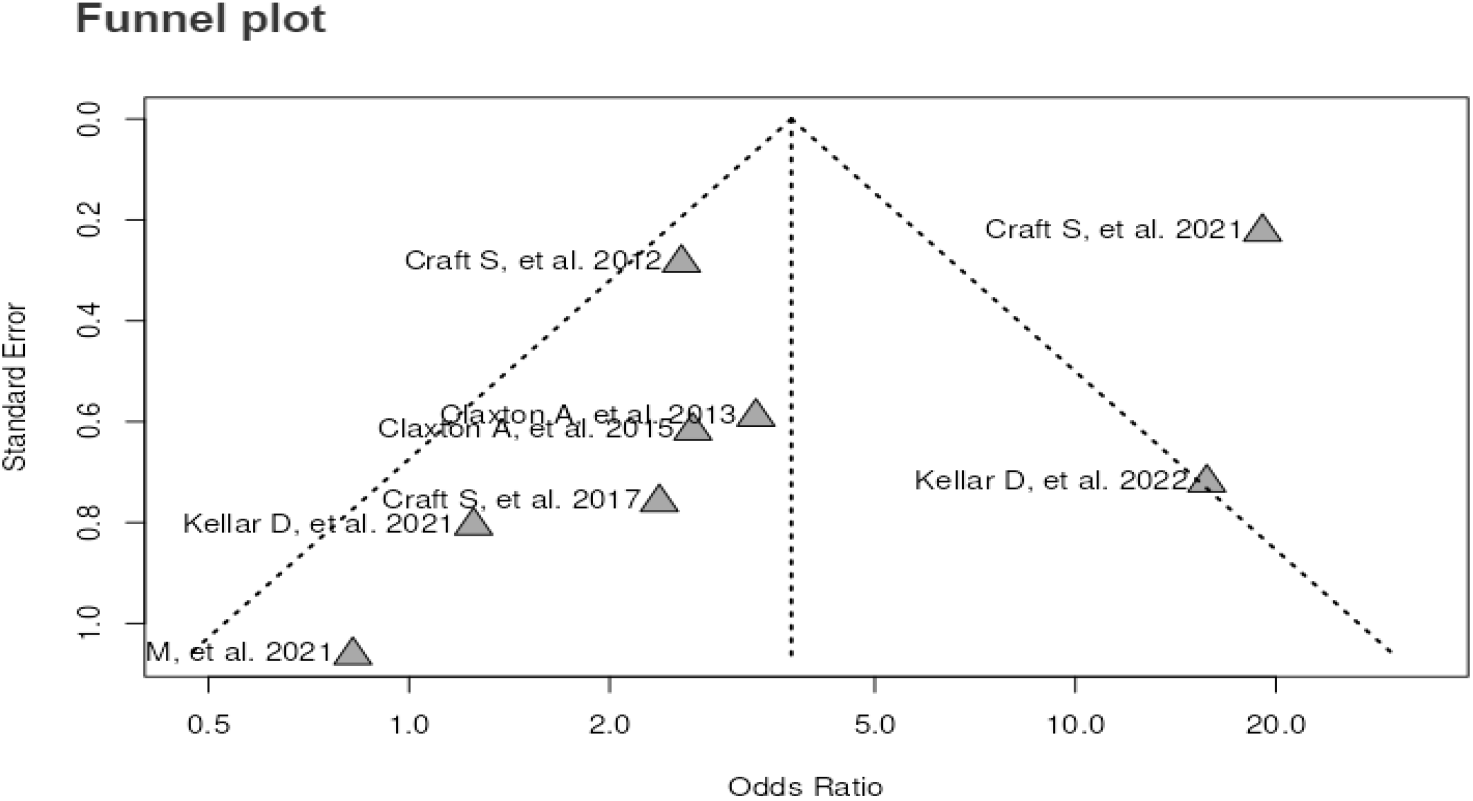
Funnel plot for meta-analysis

## DISCUSSION

The results of our systematic review and meta-analysis provide evidence that administration of INI leads to a significant improvement in cognitive function among individuals with AD compared to the placebo group. Additionally, INI demonstrates a safety profile with a low incidence of side effects.

For decades, the pancreas has been considered the primary source of insulin that traverses the blood-brain barrier. However, recent findings have brought to light the potential for insulin synthesis within the nasal epithelium and serous glands. This nasally derived insulin gains direct entry into the central nervous system via the cribriform plate and is transported along olfactory nerves to the brain parenchyma, particularly the ventromedial limbic structures.^21^. Through specific receptors, this insulin activates a cascade of insulin-dependent functions and networks within the brain, encompassing growth, metabolism, plasticity, survival, and cholinergic function, all of which are essential for learning and memory processes.^22^ A robust body of evidence has established a strong correlation between insulin receptor signaling impairments and the development of dementia, particularly AD.^23^

The identification of insulin expression and secretion capabilities within the nasal mucosa suggests a physiological route for insulin delivery to the brain. Consequently, elevating brain insulin levels has been demonstrated to enhance verbal declarative and hippocampal memory, while in AD, insulin administration improves cognitive function and decelerates cognitive decline. These observations provide compelling support for the concept of INS-based therapy for AD, encompassing both preventive and treatment strategies.^24^

The Alzheimer’s Disease Assessment Scale - Cognitive Subscale (ADAS-Cog) stands as a reliable tool for evaluating AD progression. It encompasses items assessing language, memory, praxis, and orientation, with higher scores indicating a greater degree of impairment. These items prove valuable not only in differentiating AD patients from healthy individuals but also in determining disease severity, particularly through the orientation section.^25^ Furthermore, it is important to distinguish between MCI and dementia. MCI represents a transitional stage between dementia and the age-related cognitive decline associated with normal aging, with amnestic and non-amnestic subtypes.^26^ Our meta-analysis two studies failed to identify statistically significant improvements in cognition following INI administration. However, the power to detect significant effects was limited by the small sample size and short duration of the included studies. This aligns with findings from an earlier investigation, which demonstrated that APOEε4-positive patients and female participants exhibited poorer recall following INS administration compared to non-APOEε4 carriers and male participants.^20^

The Alzheimer’s Disease Cooperative Study Activities of Daily Living Scale (ADCS-ADL) is a widely employed instrument for evaluating the functional status of individuals with AD. It assesses their ability to perform a comprehensive range of daily activities, and higher scores on the ADCS-ADL indicate a greater preservation of functional capacity, reflecting a lesser degree of impairment in daily living skills.^27^ Our investigation revealed no statistically significant disparity in ADCS-ADL scores between the insulin treatment groups and the placebo control group. However, Claxton et al.^20^ identified a sex-based discrepancy in ADCS-ADL scores, favoring females.^27^ Additionally, Craft et al. demonstrated a significant divergence in ADCS-ADL scores between the insulin and placebo groups specifically for AD patients, but not for those diagnosed with amnestic MCI.^17^

Considering the core pathophysiological processes underlying AD, a trio of protein aggregates – beta-amyloid peptides, tau protein, and hyperphosphorylated tau – is recognized as the primary culprits in AD pathogenesis.^28^ Previously, insulin was hypothesized to exert a protective effect by mitigating the accumulation of amyloid-beta peptides and reducing the phosphorylation of tau protein.^29^ However, this potential benefit is hampered by the limited ability of insulin to penetrate the blood-brain barrier.^30^ To circumvent this obstacle, intranasal insulin administration emerged as a novel therapeutic strategy. This approach capitalizes on the olfactory and trigeminal nerve pathways, allowing insulin to bypass the blood-brain barrier via perivascular transport mechanisms.^31^ Studies included in our meta-analysis evaluated the impact of INI on the cerebrospinal fluid concentrations of three important AD biomarkers: Aβ peptides, tau protein, and hyperphosphorylated tau. Interestingly, two studies by the same author reported discordant findings regarding the effects of INI administration.^17,18^ A pivotal advantage of intranasal administration lies in its ability to facilitate the direct delivery of large and charged therapeutic agents to the central nervous system via the nasal mucosa. This bypasses the blood-brain barrier, thereby minimizing systemic exposure and mitigating the potential for adverse side effects associated with various brain-targeted therapies, including those that can otherwise penetrate the blood-brain barrier.^32^ Our meta-analysis evaluated the safety profile of INI administration and revealed no statistically significant difference in the incidence of adverse events between the insulin and placebo groups. All the included studies did not report any serious adverse effects. Observed complications were limited to minor events, primarily upper respiratory symptoms and rhinitis. With the exception of a higher incidence of nasal irritation reported in Rosenbloom et al.,^15^ and a marginally higher total number of minor adverse events reported in Craft et al.,^13^ the overall findings consistently demonstrated no significant difference in complication rates between the insulin and placebo groups. Therefore, when carefully considering the risk-benefit profile of this treatment modality, INI emerges as a potentially safe therapeutic option for patients with AD.

Several antidiabetic agents have been investigated for the management of AD.^33^ INI at varying doses has been among the evaluated medications. Our meta-analysis included INI administered at doses of 20 IU or 40 IU. Among the eight studies assessed in our meta-analysis, INI at a dose of 20 IU demonstrated superior efficacy for ADAS-Cog scores in individuals with AD compared to higher doses.

The INI has been the subject of RCTs in individuals with AD. Our systematic review and meta-analysis investigating INI as a treatment for AD in the context of in situ diabetes was constrained by several limitations, including a relatively small overall sample size, heterogeneity in the number of participants across studies (ranging from 12 to 121), variability in study duration (from 4 to 48 months), and differences in the types and dosages of insulin employed. Despite these limitations, our findings provide robust evidence supporting the safety and efficacy of INI in this patient population.

## CONCLUSION

This systematic review and meta-analysis evaluated the effects of INI administration in individuals with AD, and while the data is not yet definitive enough to establish INI as a definitive treatment for AD, the accumulating evidence supporting its safety, efficacy, and reduced systemic side effects strongly suggests that INI is associated with an overall enhancement of global cognition. The study’s findings demonstrated a significant improvement in ADAS-Cog scores among participants, particularly those receiving a 20 IU INI dosage. Furthermore, the observed adverse effects were minimal. Given the nascent nature of this research field, further studies are warranted to elucidate the heterogeneity in treatment response to INI and extend its pro-cognitive benefits to broader patient populations, aiming to improve the quality of life of individuals with AD.

## Data Availability

All data produced in the present work are contained in the manuscript

